# Development and validation of automated computer aided-risk score for predicting the risk of in-hospital mortality using first electronically recorded blood test results and vital signs for COVID-19 hospital admissions: a retrospective development and validation study

**DOI:** 10.1101/2020.11.30.20241273

**Authors:** Muhammad Faisal, Mohammed A Mohammed, Donald Richardson, Massimo Fiori, Kevin Beatson

## Abstract

**Objectives:** There are no established mortality risk equations specifically for unplanned emergency medical admissions which include patients with the novel coronavirus SARS-19 (COVID-19). We aim to develop and validate a computer-aided risk score (CARMc19) for predicting mortality risk by combining COVID-19 status, the first electronically recorded blood test results and latest version of the National Early Warning Score (NEWS2).

**Design:** Logistic regression model development and validation study using a cohort of unplanned emergency medical admissions to hospital.

**Setting:** York Hospital (YH) as model development dataset and Scarborough Hospital (SH) as model validation dataset.

**Participants:** Unplanned adult medical admissions discharged over three months (11 March 2020 to 13 June 2020) from two hospitals (YH for model development; SH for external model validation) based on admission NEWS2 electronically recorded within ±24 hours and/or blood test results within ±96 hours of admission. We used logistic regression modelling to predict the risk of in-hospital mortality using two models: 1) CARMc19_N: age + sex + NEWS2 including subcomponents + COVID19; 2) CARMc19_NB: CARMc19_N in conjunction with seven blood test results and acute kidney injury score. Model performance was evaluated according to discrimination (c-statistic), calibration (graphically), and clinical usefulness at NEWS2 thresholds of 4+, 5+, 6+.

**Results:** The risk of in-hospital mortality following emergency medical admission was similar in development and validation datasets (8.4% vs 8.2%). The c-statistics for predicting mortality for Model CARMc19_NB is better than CARMc19_N in the validation dataset (CARMc19_NB = 0.88 (95%CI 0.86 to 0.90) vs CARMc19_N = 0.86 (95%CI 0.83 to 0.88)). Both models had good internal and external calibration (CARMc19_NB: 1.01 (95%CI 0.88 vs 1.14) & CARMc19_N: 0.95 (95%CI 0.83 to 1.06)). At all NEWS2 thresholds (4+, 5+, 6+) model CARMc19_NB had better sensitivity and similar specificity.

**Conclusions:** We have developed a validated CARMc19 score with good performance characteristics for predicting the risk of in-hospital mortality following an emergency medical admission using the patient’s first, electronically recorded vital signs and blood tests results. Since the CARMc19 scores place no additional data collection burden on clinicians and is readily automated, it may now be carefully introduced and evaluated in hospitals with sufficient informatics infrastructure.

## Introduction

The novel coronavirus SARS-19 produces the newly identified disease ‘COVID-19’ in patients with symptoms (Coronaviridae Study Group of the International Committee on Taxonomy of Viruses[1]) and was declared a pandemic on 11 March 2020 that has challenged health care systems worldwide.

COVID-19 patients admitted to hospital can develop severe disease with life threatening respiratory and/or multi-organ failure [2,3] with a high risk of mortality. The appropriate early assessment and management of patients with COVID-19 is important in ensuring high-quality care including isolation, escalation to critical care or palliative care.

Early Warning Scores (EWS) are commonly used in hospitals worldwide [4], and in the National Health Service (NHS) hospitals in England, the patient’s vital signs are monitored and summarised into a National Early Warning Score (NEWS)[5]. We have developed two automated risk equations to predict the patient’s risk of mortality (CARM_N & CARM_NB) using NEWS only (CARM_N) [6] and NEWS + blood test results (CARM_NB) [7] following emergency medical admission to hospital. We found CARM performed similar to consultant clinicians [8].

In December 2017, an update to NEWS (NEWS2) was published [4] that extends the level of consciousness from AVPU to ACVPU, where C represents new confusion or delirium and is allocated three points (the maximum for a single variable). NEWS2 also offers two scales for oxygen saturation (scale 1 and scale 2) which accommodates patients with hypercapnic respiratory failure who have clinically recommended oxygen saturation of 88–92%.

Whilst hospitals continue to use NEWS2 during the COVID-19 pandemic, little is known about how NEWS2 and CARM scores perform in monitoring COVID-19 patients. In this study, we aimed to develop and validate an automated computer aided risk score (CARMc19) using on admission NEWS2 and blood test results for predicting mortality. This approach is clinically useful because it places no additional data collection burden on staff for monitoring COVID-19 patients.

## Methods

### Setting & data

Our cohorts of emergency medical admissions are from two acute hospitals which are approximately 65 kilometres apart in the Yorkshire & Humberside region of England – Scarborough hospital (n∼300 beds) and York Hospital (YH) (n∼700 beds), managed by York Teaching Hospitals NHS Foundation Trust. We selected these hospitals because they had electronic NEWS2, collected as part of the patient’s process of care since April 2019, and were agreeable to the study.

We considered all consecutive adult (age≥18 years) non-elective or emergency medical admissions discharged over a course of three months (11 March 2020 to 13 June 2020) with electronic NEWS2. For each emergency admission, we obtained a pseudonymised patient identifier, patient’s age (years), sex (male/female), discharge status (alive/dead), admission and discharge date and time, diagnoses codes based on the 10th revision of the International Statistical Classification of Diseases (ICD-10), NEWS2 (including its subcomponents respiratory rate, temperature, systolic pressure, pulse rate, oxygen saturation, oxygen supplementation, oxygen scales 1 & 2, and alertness including confusion), blood test results (albumin, creatinine, haemoglobin, potassium, sodium, urea, and white cell count), and Acute Kidney Injury (AKI) score.

The diastolic blood pressure was recorded at the same time as systolic blood pressure. Historically, diastolic blood pressure has always been a routinely collected physiological variable on vital sign charts and is still collected where electronic observations are in place. NEWS2 produces integer values that range from 0 (indicating the lowest severity of illness) to 20 (the maximum NEWS2 value possible) (see Table S1 in supplementary material). The index NEWS2 was defined as the first electronically recorded NEWS2 within ±24 hours of the admission time. We excluded records where the index NEWS2 (or blood test results) was not within ±24 hours (±96 hours) or was missing/not recorded at all (see Table S2). The ICD-10 code ‘U071’ was used to identify records with COVID-19. We searched primary and secondary ICD-10 codes for ‘U071’ for identifying COVID-19.

## Statistical Modelling

We began with exploratory analyses including box plots and line plots to show the relationship between covariates and risk of in-hospital mortality. We developed two logistic regression models, known as CARMc19_N and CARMc19_NB, to predict the risk of in-hospital mortality with following covariates: 1) Model CARMc19_N uses age + sex + COVID-19 (yes/no) + NEWS2 including sub components; 2) Model CARMc19_NB extends Model CARMc19_N with all seven blood test results and AKI score. The primary rationale for using these variables is that they are routinely collected as part of process of care and their inclusion in our statistical models is on clinical grounds as opposed to the statistical significance of any given covariate.

We used the qladder function (Stata [9]), which displays the quantiles of a transformed variable against the quantiles of a normal distribution according to the ladder powers 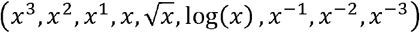 for each continuous covariate and chose the following transformations: (creatinine)-1/2, log_e_(potassium), log_e_(white cell count), log_e_(urea), log_e_ (respiratory rate), log_e_(pulse rate), log_e_(systolic blood pressure), and log_e_(diastolic blood pressure). We used an automated approach to search for all two-way interactions and incorporated those interactions which were statistically significant (p<0.001) from the MASS library [12] in *R* [13].

We developed both models using York Hospital (YH) data (development dataset) and externally validated their performance on Scarborough Hospital (SH) data (validation dataset). The hospitals are part of the same NHS Trust but are geographically separated by about 65 kilometres (40 miles).

We report discrimination and calibration statistics as performance measures for these models [10].

Discrimination relates to how well a model can separate – or discriminate between – those who died and those who did not and is given by the area under the Receiver Operating Characteristics (ROC) curve (AUC) or c-statistic. The ROC curve is a plot of the sensitivity (true positive rate) versus 1-specificity (false positive rate) for consecutive predicted risks. A c-statistic of 0.5 is no better than tossing a coin, whilst a perfect model has a c-statistic of 1. In general, values less than 0.7 are considered to show poor discrimination, values of 0.7 to 0.8 can be described as reasonable, and values above 0.8 suggest good discrimination [11]. The 95% confidence interval for the c-statistic was derived using DeLong’s method as implemented in the *pROC* library [12] in R [13].

Calibration measures a model’s ability to generate predictions that are, on average, close to the average observed outcome and can be readily seen on a scatter plot (y-axis = observed risk, x-axis = predicted risk). Perfect predictions should be on the 45° line. We internally validated and assessed the calibration for all the models using the bootstrapping approach [14,15]. The overall statistical performance was assessed using the scaled Brier score which incorporates both discrimination and calibration [10]. The Brier score is the squared difference between actual outcomes and predicted risk of death, scaled by the maximum Brier score such that the scaled Brier score ranges from 0– 100%. Higher values indicate superior models.

The recommended threshold for detecting deteriorating patients and sepsis is NEWS2≥5 [16,17]. Therefore, we assessed the sensitivity, specificity, positive and negative predictive values and likelihood ratios for these models at NEWS2 threshold of 4+,5+, and 6+ [18]. We followed the TRIPOD guidelines for reporting of model development and validation [19]. We used Stata [9] for data cleaning and R [13] for statistical analysis.

## Results

### Cohort Characteristics

The number of non-elective discharges was 6444 over 3 months. For the development of CARMc19_N, we excluded 36 (0.6%) of admissions because the index NEWS2 was not recorded within ±24 hours of the admission date/time, or they were missing or not recorded at all (see Table S2). Likewise, for the development of CARMc19_NB, we further excluded 1189 (18.3%) of admissions because the first blood test results were not recorded within ±96 hours of the admission date/time, or they were missing or not recorded at all (see Table S2).

The characteristics of the admissions included in our study are shown in Table 1. Emergency admissions in the validation dataset were older than those in development dataset (69.6 years vs 67.4 years), less likely to be male (49.5% vs 51.2%), had higher index NEWS2 (3.2 vs 2.8), higher prevalence of COVID-19 (11.0% vs 8.7%) but similar in-hospital mortality (8.4% vs 8.2%). See accompanying scatter and boxplots in Figure S1 to S4 (supplemental digital content).

**Table 1.**
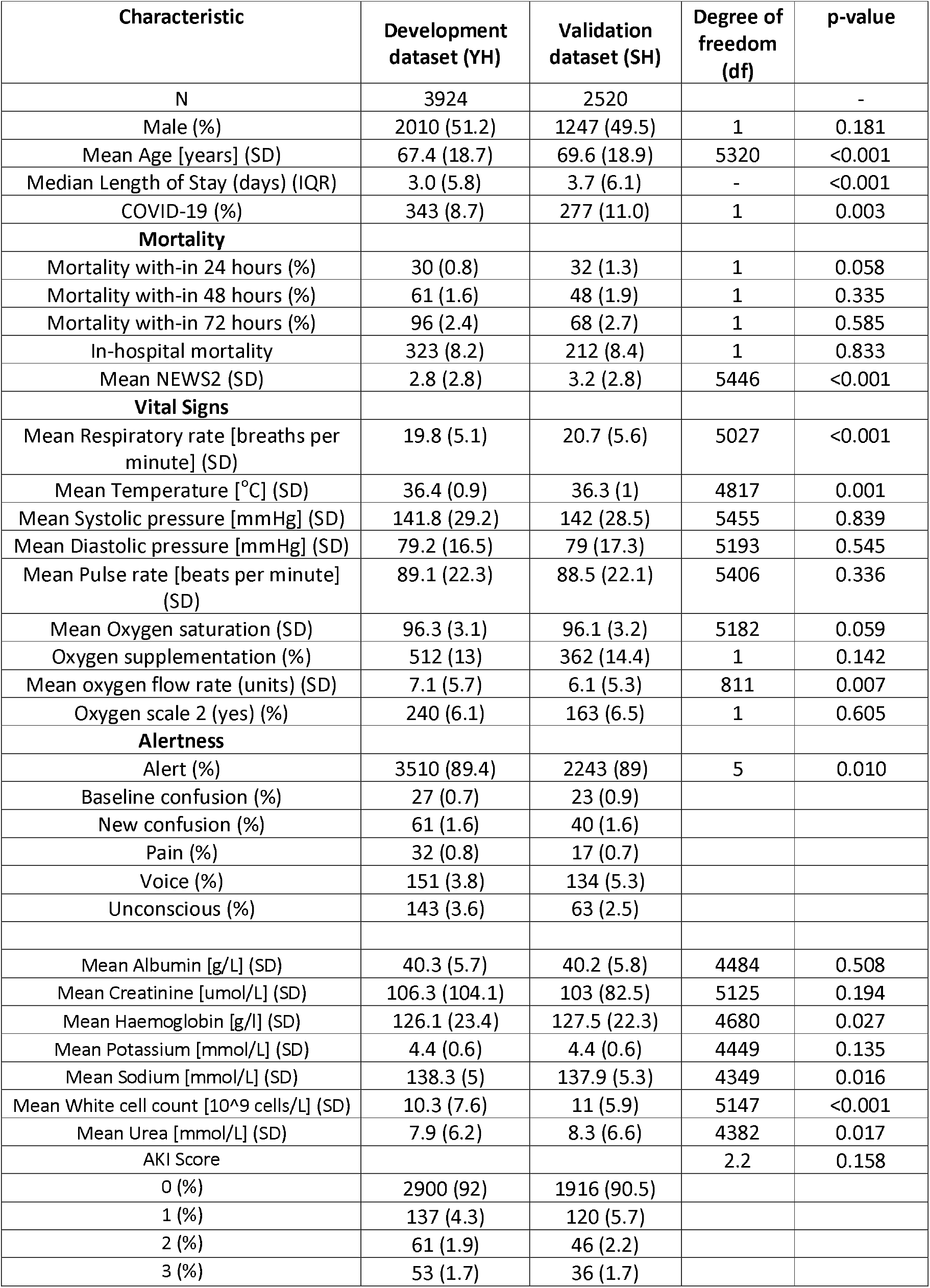
**Characteristics of emergency medical admissions in development and validation datasets**.

We assessed the performance of both (CARMc19_N and CARMc19_NB) models to predict the risk of in-hospital mortality in emergency medical admissions (see Table 2 and Figure 1 for validation results and Table S3 and Figure S7 for model development results).

**Table 2:**
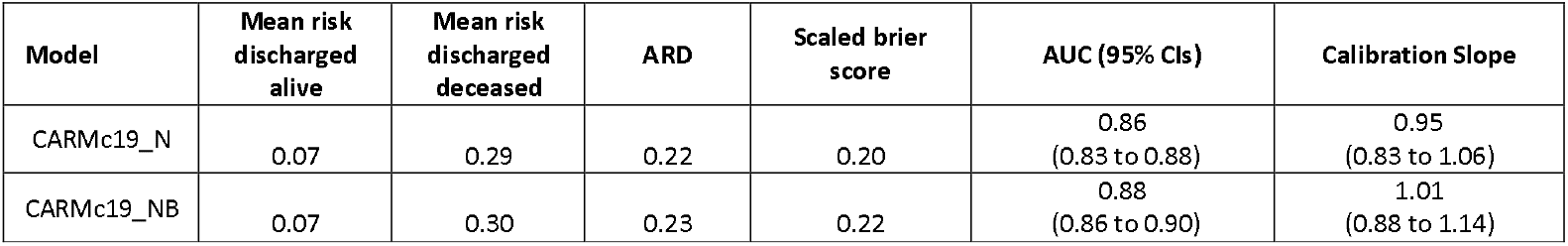
**Performance of CARMc19_N and CARMc19_NB models for predicting the risk of mortality in validation dataset**

**Figure 1.**
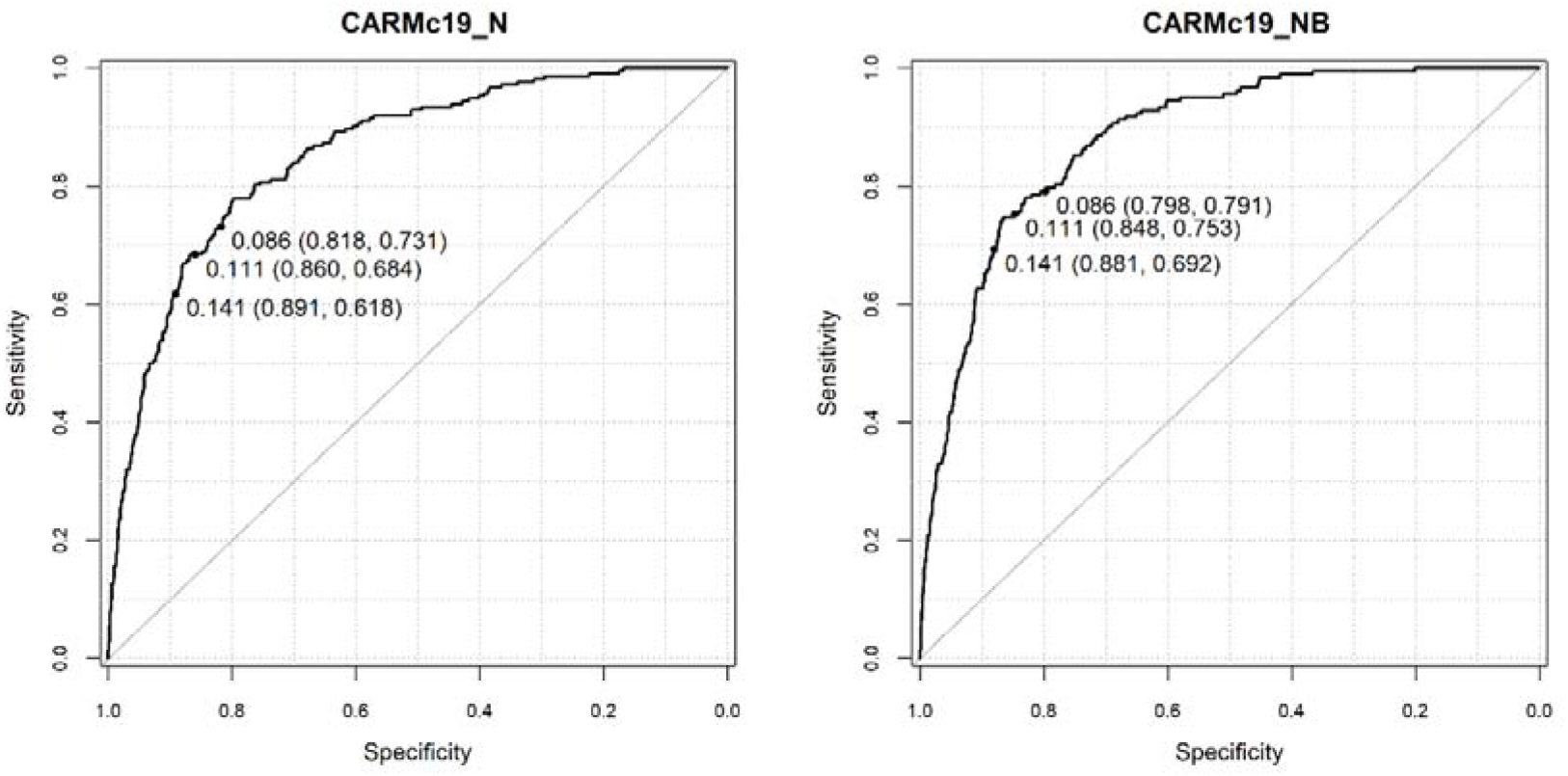
Receiver Operating Characteristic curve for CARMc19_N and CARMc19_NB in predicting the risk of mortality in the development dataset. Note: predicted probability at NEWS2 thresholds 4+ [0.09], 5+ [0.11], 6+ [0.14] (sensitivity, specificity)

The c-statistics for predicting mortality for Model CARMc19_NB is better than Model CARMc19_N in development dataset (CARMc19_NB = 0.87 (95%CI 0.85 to 0.89) vs CARMc19_N = 0.86 (95%CI 0.84 to 0.87)) and the validation dataset (CARMc19_NB = 0.88 (95%CI 0.86 to 0.90) vs CARMc19_N = 0.86 (95%CI 0.83 to 0.88)).

Internal validation of both models is shown in Figure S6. Both models had good internal and external calibration (CARMc19_NB: 1.01 (95%CI 0.88 vs 1.14) & CARMc19_N: 0.95 (95%CI 0.83 to 1.06)) (see Table 2 and Figure 2).

**Figure 2.**
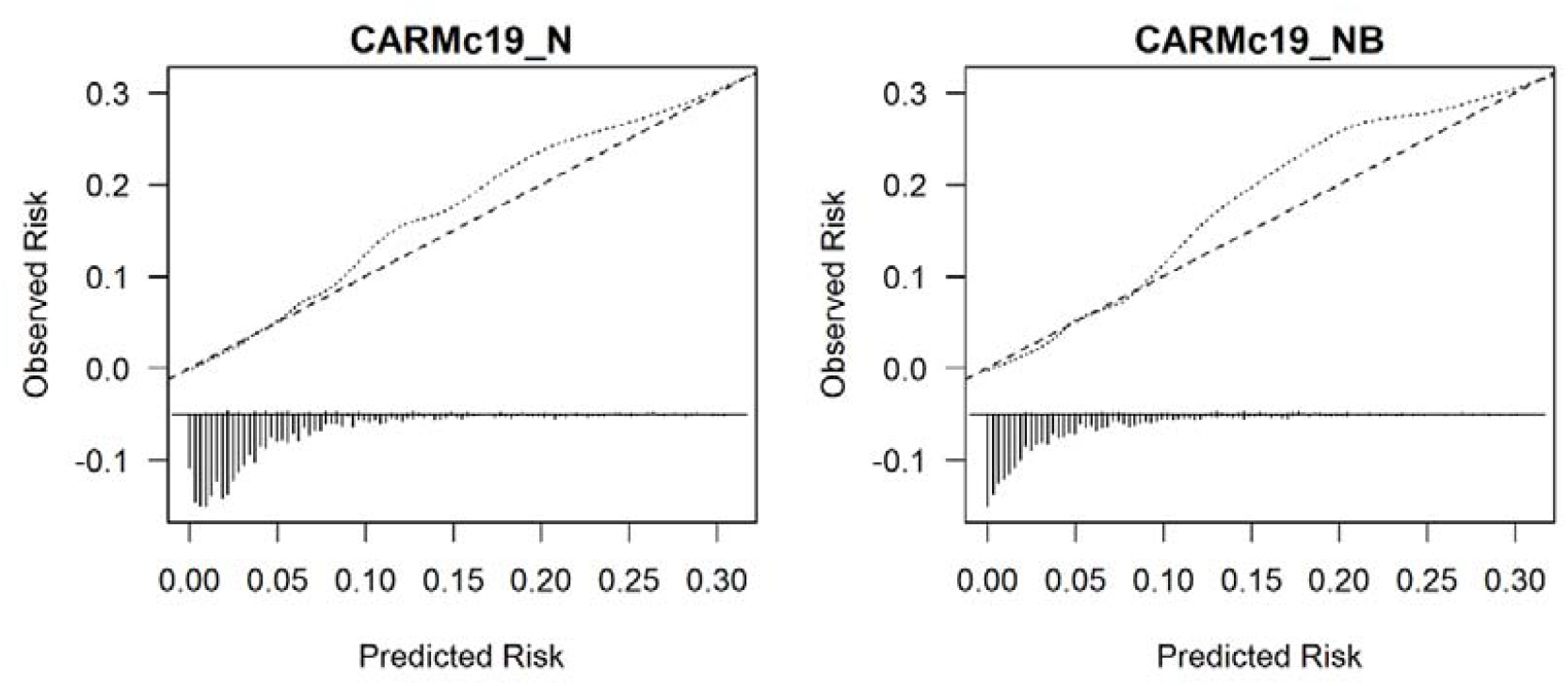
External validation of CARMc19_N and CARMc19_NB models, respectively for predicting the risk of mortality. NB: We limit the risk of mortality to 0.30 for visualisation purpose because beyond this point, we have few patients.

Table 3 includes the sensitivity, specificity, positive and negative predictive values for CARMc19_N and CARMc19_NB models for predicting mortality at NEWS2 threshold of 4+, 5+, 6+.

**Table 3.** **Sensitivity analysis of CARMc19_N and CARMc19_NB models in validation dataset for predicting the risk of mortality at NEWS2 thresholds 4+ [0**.**09]**, **5+ [0**.**11], and 6+ [0**.**14] of predicted risk of mortality in development dataset**.

| Model | At NEWS score [predicted risk of death] | Number of deaths identified by model | Sensitivity% | Specificity% | PPV | NPV | LR+ | LR- |
| --- | --- | --- | --- | --- | --- | --- | --- | --- |
| CARMc19_N | 4+ [0.09] | 696 | 73.1<br>(66.6 to 79) | 81.8<br>(80.2 to 83.4) | 27<br>(23.4 to 30.8) | 97.1<br>(96.2 to 97.8) | 4<br>(3.6 to 4.5) | 0.3<br>(0.3 to 0.4) |
| CARMc19_N | 5+ [0.11] | 557 | 68.4<br>(61.7 to 74.6) | 86<br>(84.5 to 87.4) | 31<br>(26.8 to 35.4) | 96.7<br>(95.9 to 97.5) | 4.9<br>(4.3 to 5.6) | 0.4<br>(0.3 to 0.4) |
| CARMc19_N | 6+ [0.14] | 452 | 61.8<br>(54.9 to 68.4) | 89.1<br>(87.8 to 90.4) | 34.3<br>(29.5 to 39.3) | 96.2<br>(95.3 to 97) | 5.7<br>(4.9 to 6.7) | 0.4<br>(0.4 to 0.5) |
| CARMc19_NB | 4+ [0.09] | 651 | 79.1<br>(72.5 to 84.8) | 79.8<br>(77.9 to 81.6) | 26.9<br>(23.2 to 30.9) | 97.6<br>(96.7 to 98.3) | 3.9<br>(3.5 to 4.4) | 0.3<br>(0.2 to 0.3) |
| CARMc19_NB | 5+ [0.11] | 526 | 75.3<br>(68.3 to 81.4) | 84.8<br>(83.1 to 86.3) | 31.7<br>(27.3 to 36.3) | 97.3<br>(96.4 to 98) | 4.9<br>(4.3 to 5.6) | 0.3<br>(0.2 to 0.4) |
| CARMc19_NB | 6+ [0.14] | 431 | 69.2<br>(62 to 75.8) | 88.1<br>(86.5 to 89.5) | 35.3<br>(30.3 to 40.5) | 96.8<br>(95.9 to 97.6) | 5.8<br>(5 to 6.8) | 0.3<br>(0.3 to 0.4) |

PPV = Positive Predictive Value; NPV = Negative Predictive Value; LR+ = Positive Likelihood Ratio; LR- = Negative Likelihood Ratio

At all NEWS2 thresholds (4+, 5+, 6+) model CARMc19_NB had better sensitivity (development dataset: 76% vs 72%; 71% vs 67%; 65% vs 61% and validation dataset: 79% vs 73%; 75% vs 68%; 69% vs 61%) and similar specificity (development dataset: 81% vs 82%; 86% vs 86%; 89% vs 90% and validation dataset: 80% vs 82%; 85% vs 86%; 88% vs 89%) (see Table 3 & S4).

## Discussion

In this study, we developed and validated two (CARMc19_N and CARMc19_NB) models to predict the risk of in-hospital mortality with the following covariates: 1) Model CARMc19_N uses age + sex + COVID-19 (yes/no) + NEWS2 including sub components; 2) Model CARMc19_NB extends Model CARMc19_N with all seven blood test results and AKI score (see appendix for equations and escalation policy Figure S5). We found that CARMc19 scores has good performance and our findings tentatively suggest that a NEWS2 threshold of 5+ appears to strike a reasonable balance between sensitivity and specificity. Model CARMc19_NB was more sensitive with similar specificity than the CARMc19_N model.

CARMc19 scores performed better than our previous CARM models [6] [7] because of additional NEWS2 variables (oxygen flow rate & oxygen scale 2) and COVID-19 status. A recent systematic review identified models to predict mortality from COVID-19 with c-statistics that ranged from 0.87 to 1 [20]. However, despite these high c-statistics, the review authors cautioned against the use of these models in clinical practice because of the high risk of bias and poor reporting of studies which are likely to have led to optimistic results [20].

The main advantages of our models are that they are designed to incorporate data which are already available in the patient’s electronic health record thus placing no additional data collection or computational burden on clinicians, and are readily automated. Nonetheless, we emphasise that our computer-aided risk scores are not designed to replace clinical judgement. They are intended and designed to support, not subvert, the clinical decision-making process and can be always overridden by clinical concern [5,21]. The working hypothesis for our models is that they may enhance situational awareness of mortality by processing information already available without impeding the workflow of clinical staff, especially as our approach offers a faster and less expensive assessment of in-hospital mortality risk than current laboratory tests which may be more practical to use for large numbers of people.

There are limitations in relation to our study. We identified COVID-19 based on ICD-10 code ‘U071’ which was determined by COVID-19 swab test results (hospital or community) and clinical judgment and so our findings are constrained by the accuracy of these methods [22,23]. This does, however, allow the algorithm to take account of the entry of diagnostic information by the clinician including radiology findings as input variables if the swab result is negative. We used the index NEWS2 data in our models, but vital signs and blood test results are repeatedly updated for each patient according to hospital protocols. Although we developed models using one hospital’s data and validated into another hospital’s data, the extent to which changes in vital signs over time reflect changes in mortality risk need to be incorporated in our models requires further study. Our two hospitals are part of the same NHS Trust and this may undermine the generalisability of our findings, which merit further external validation.

Although we focused on in-hospital mortality (because we aimed to aid clinical decision making in the hospital), the impact of this selection bias needs to be assessed by capturing out-of-hospital mortality by linking death certification data and hospital data. CARMc19, like other risk scores, can only be an aid to the decision-making process of clinical teams [11,24] and its usefulness in clinical practice remains to be seen.

The next phase of this work is to field test CARMc19 scores by carefully engineering it into routine clinical practice to see if it does enhance the quality of care for acutely ill patients, whilst noting any unintended consequences.

## Conclusion

We have developed a validated CARMc19 score with good performance characteristics for predicting the risk of in-hospital mortality following an emergency medical admission using the patient’s first, electronically recorded, vital signs and blood tests results. Since the CARMc19 scores place no additional data collection burden on clinicians and is readily automated, it may now be carefully introduced and evaluated in hospitals with sufficient informatics infrastructure.

## Data Availability

Our data sharing agreement is with York hospital and does not permit us to share the data used in this paper.

## Author Contributions

DR and MAM had the original idea for the work. KB, RH provided the data extracts. MF undertook the statistical analyses with support from MAM. MF, MAM, and DR wrote the first draft of the paper. DR provided clinical perspectives. All others contributed to the final paper and have approved the final version. DR & MF will act as study guarantors.

## Transparency Declaration

The lead author (the manuscript’s guarantor) affirms that the manuscript is an honest, accurate, and transparent account of the study being reported and that no important aspects of the study have been omitted.

## Ethical Approval

This study was deemed to be exempt from ethical approval because it was classified as an evaluation. Furthermore, this study used already de-identified data from an ongoing study involving NEWS which received ethical approval from Health Research Authority (HRA) and Health and Care Research Wales (HCRW) (reference number 19/HRA/0548).

## Patient Involvement

No1ne.

