## supplementary material for "Development and validation of automated computer aided-risk score for predicting the risk of in-hospital mortality using first electronically recorded blood test results and vital signs for COVID-19 hospital admissions: a retrospective development and validation study"

### **Model CARMc19_N:** predicting in-hospital mortality using vital signs only (N)

$$\boldsymbol{Logit}\left( \boldsymbol{Died} \right)\boldsymbol{= 10.671+ 9.262 * Male + 0.044 * Age+ 0.069 * NEWS}\boldsymbol{2+ 0.845 * log}\left( \boldsymbol{Respiratory Rate} \right)\boldsymbol{- 0.276 * Temperature - 0.022 * log}\left( \boldsymbol{Diastolic pressure} \right)\boldsymbol{-1.524* log}\left( \boldsymbol{Systolic pressure} \right)\boldsymbol{+0.438 * log}\left( \boldsymbol{Pulse rate} \right)\boldsymbol{- 0.044 * Oxygen Saturations+ 0.125 * Oxygen Supplementation+0.487*Baseline Confusion-19.329*New Confusion+ 0.789 *Pain + 0.476* Voice+1.608*Unconscious+0.349*Oxygen Scale 2+0.056*Oxygen Flow Rate-8.947*COVID}\boldsymbol{19-0.096*Male*Oxygen Saturation+3.856*log}\left( \boldsymbol{Systolic pressure} \right)\boldsymbol{*New Confusion+0.085*Oxygen Saturation*COVID}\boldsymbol{19+0.036*Age*COVID}\boldsymbol{19}$$

### **Model CARMc19_NB:** predicting in-hospital mortality using vital signs (N) and blood test results (B)

$$\boldsymbol{Logit}\left( \boldsymbol{Died} \right)\boldsymbol{= 14.241 -1.478*Male +0.041*Age -0.063*Albumin} \boldsymbol{+5.900*log}\left( \boldsymbol{Creatinine} \right)\boldsymbol{-0.002*Haemoglobin -0.830*log}\left( \boldsymbol{Potassium} \right)\boldsymbol{-0.043*Sodium +0.342*log}\left( \boldsymbol{White Cell Count} \right)\boldsymbol{+0.181*log}\left( \boldsymbol{Urea} \right)\boldsymbol{+0.434*AKI Score 1 +0.465*AKI Score 2 +0.406*AKI Score 3 +0.050*NEWS}\boldsymbol{2 +0.729*log}\left( \boldsymbol{Respiratory Rate} \right)\boldsymbol{-0.257*Temperature+0.863*log}\left( \boldsymbol{Diastolic pressure} \right)\boldsymbol{-0.875* log}\left( \boldsymbol{Systolic pressure} \right)\boldsymbol{+0.066*log}\left( \boldsymbol{Pulse rate} \right)\boldsymbol{-0.055*Oxygen saturation +0.175*Oxygen supplementation +0.393*Baseline Confusion -0.414*New Confusion +1.085* Pain +1.024* Voice +2.635*Unconscious+0.611*Oxygen Scale 2 +0.026*Oxygen Flow rate +1.934*COVID}\boldsymbol{19 +0.648*Male*log}\left( \boldsymbol{Urea} \right)\boldsymbol{+0.127*New Confusion*log(White Cell Count)}$$

**We accounted for baseline difference in risk of mortality in the external validation data by subtracting (CARMc19_N: 0.24 and CARMc19_NB:0.27) from the CARMc19 logit models using an iterative procedure described elsewhere^1^.**

**1.	Faisal M, Howes R, Steyerberg EW, Richardson D, Mohammed MA. Using routine blood test results to predict the risk of death for emergency medical admissions to hospital: an external model validation study. QJM [Internet]. 2017 Jan 1 [cited 2017 Oct 2];110(1):27–31. Available from: https://academic.oup.com/qjmed/article-lookup/doi/10.1093/qjmed/hcw110**

**Table S1: NEWS2 scoring chart**

| **Physiological Parameters** | **3** | **2** | **1** | **0** | **1** | **2** | **3** |
| --- | --- | --- | --- | --- | --- | --- | --- |
| **Respiration Rate** | **≤8** |  | **9 - 11** | **12 - 20** |  | **21 - 24** | **≥25** |
| **SpO2 Scale 1 (%)** | **≤91** | **92 - 93** | **94 - 95** | **≥96** |  |  |  |
| **SpO2 Scale 2 (%)** | **≤83** | **84 - 85** | **86 - 87** | **88 - 92**  **≥93 on Air** | **93 – 94 on oxygen** | **95 – 96 on oxygen** | **≥97 on oxygen** |
| **Oxygen Saturations** | **≤91** | **92 - 93** | **94 - 95** | **≥96** |  |  |  |
| **Air or Oxygen?** |  | **Oxygen** |  | **Air** |  |  |  |
| **Temperature** | **≤35.0** |  | **35.1 - 36.0** | **36.1 - 38.0** | **38.1 - 39.0** | **≥39.1** |  |
| **Systolic BP** | **≤90** | **91 - 100** | **101 - 110** | **111 - 219** |  |  | **≥220** |
| **Heart Rate** | **≤40** |  | **41 - 50** | **51-90** | **91 - 110** | **111 - 130** | **≥131** |
| **Level of Consciousness** |  |  |  | **Alert** |  |  | **Voice, Pain, Confusion, or Unconscious** |

**The NEWS [https://www.rcplondon.ac.uk/projects/outputs/national-early-warning-score-news] is based on a scoring system in which a score is allocated to the physiological measurements of vital signs already undertaken when patients present to or are being monitored in hospital. A score is allocated to each as they are measured, the magnitude of the score reflecting how extreme the parameter varies from the norm. This score is then aggregated and uplifted for people requiring oxygen.**

| **Characteristic** | **Development dataset (YH)** | **Validation dataset (SH)** | **All** |
| --- | --- | --- | --- |
|  | **N (%)** | **N (%)** | **N (%)** |
| **Total emergency medical discharges between**  **11 Mar 20 to 13 June 20** | 3952 | 2528 | 6480 |
| **Excluded: No NEWS2 recorded (%)** | 13 (0.3) | 6 (0.2) | 19 (0.3) |
| **Excluded: First NEWS2 after 24 hours of admission (%)** | 15 (0.4) | 2 (0.1) | 17 (0.3) |
| **Excluded: First blood test results after 4 days of admission (%)** | 786 (20.0) | 403 (15.9) | 1189 (18.3) |
| **Total excluded (%)** | 814 (20.6) | 411 (16.3) | 1225 (18.9) |
| **Total included (%)** | 3138 (79.4) | 2117 (84.7) | 5255 (81.1) |

**Table S2 Number of emergency medical admissions included/excluded**

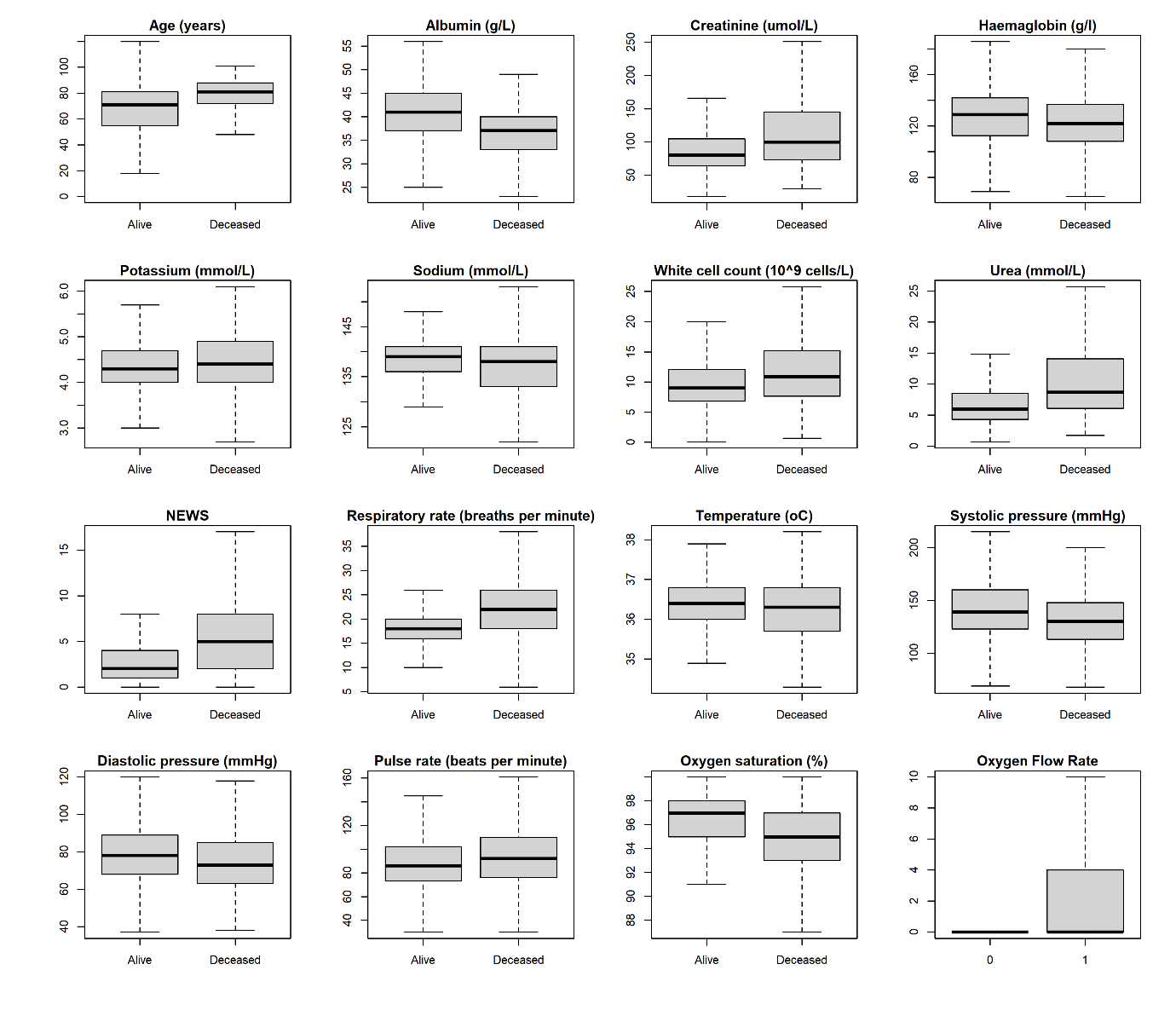

#### Figure S1 Boxplot for continuous covariates without outliers with respect to discharge status (alive/deceased) for development dataset.

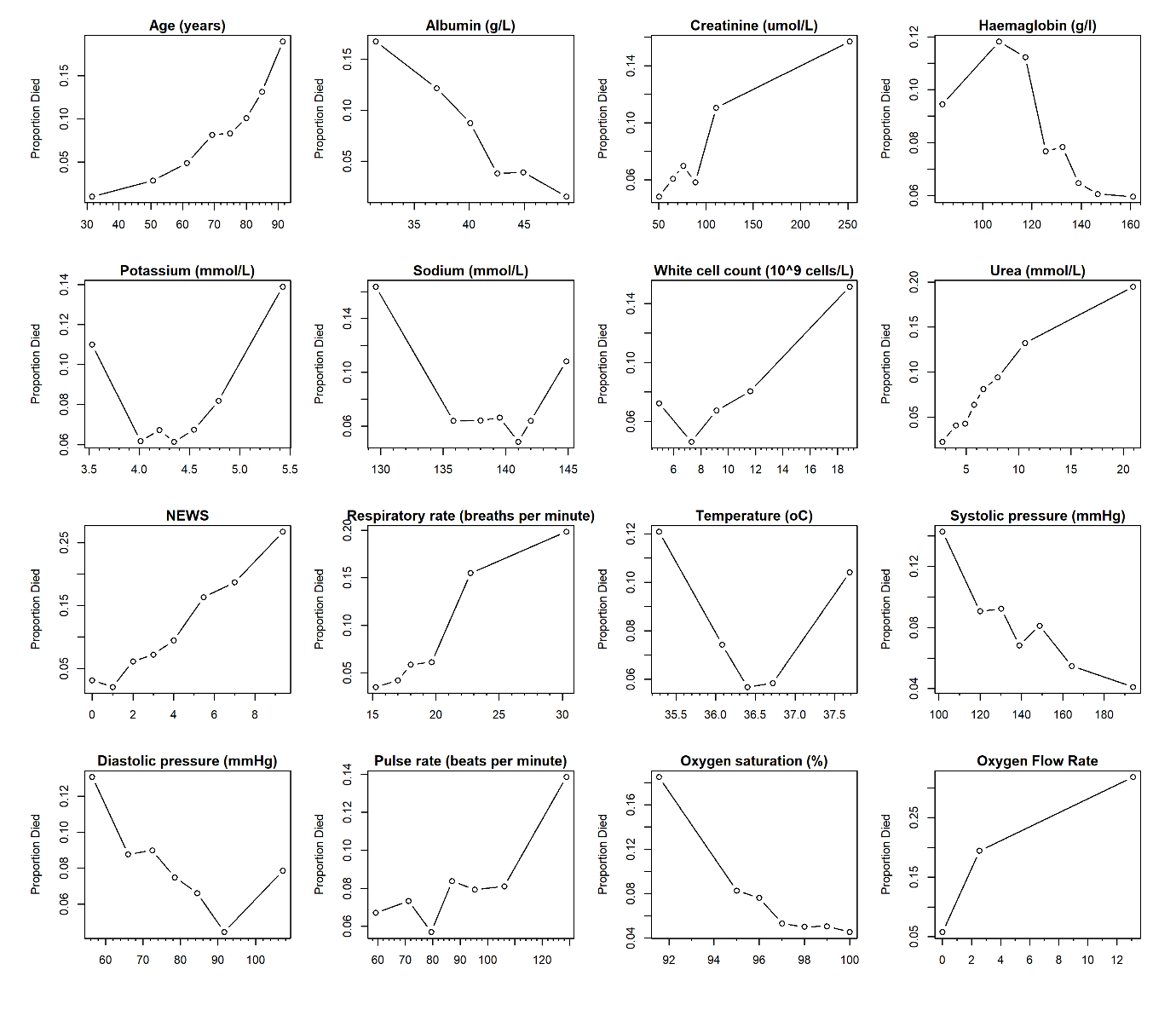

Figure S2 Scatter plots showing the observed risk of in-hospital mortality with continuous covariates for development dataset.

NB: y-axis range changes in each plot.

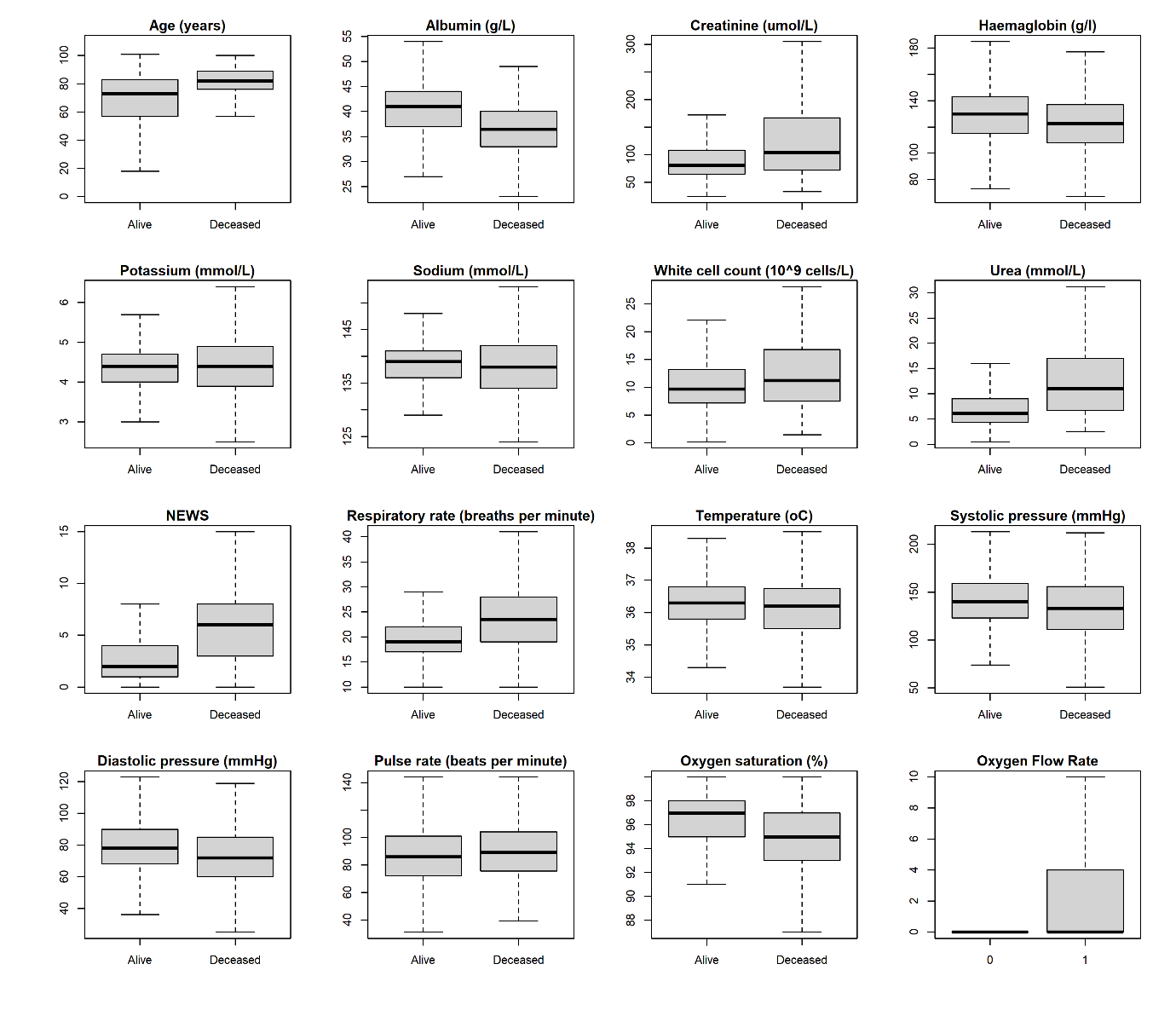

Figure S3 Boxplot for continuous covariates without outliers with respect to discharge status (alive/deceased) for validation dataset.

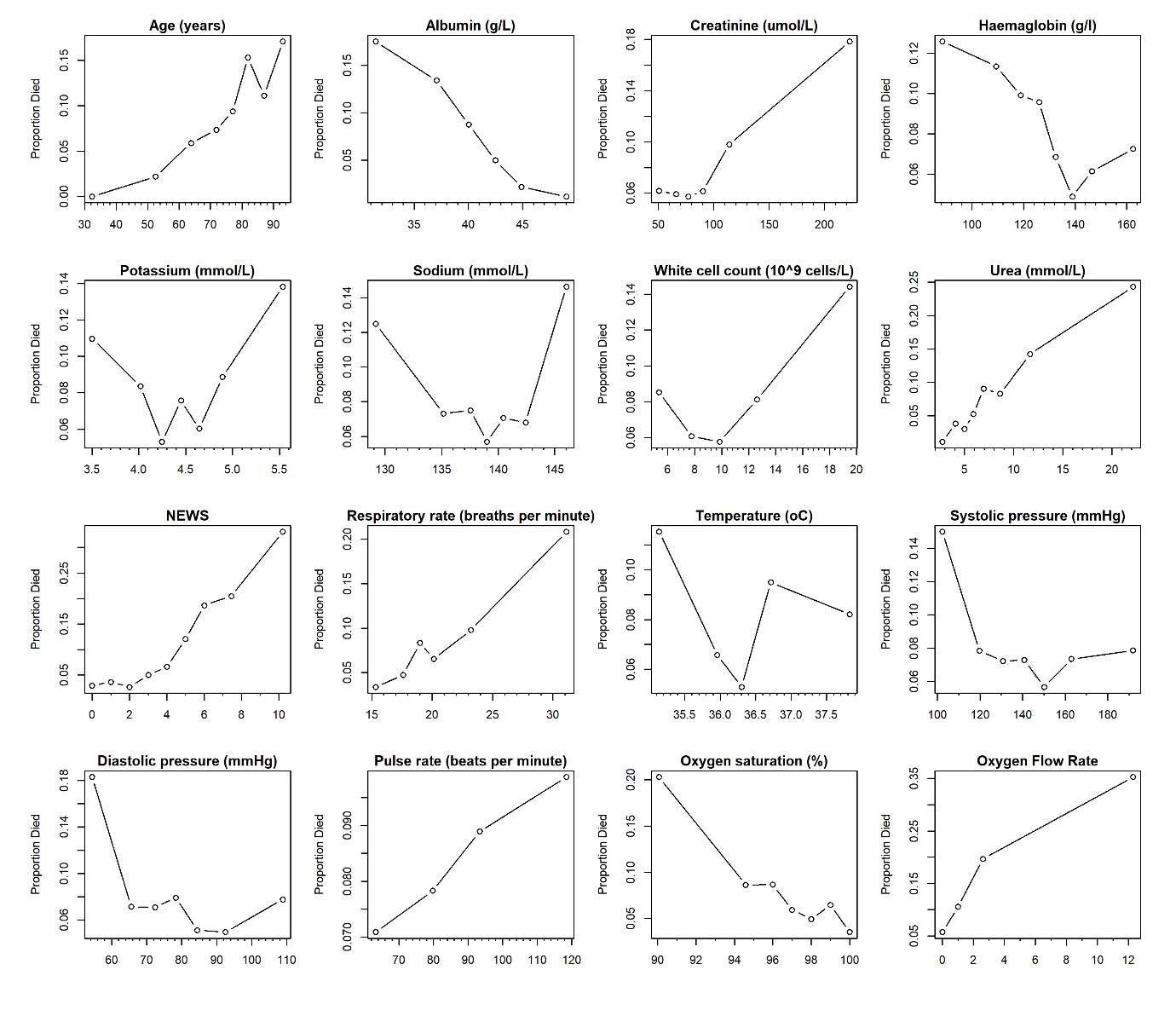

Figure S4 Scatter plots showing the observed risk of in-hospital mortality with continuous covariates for validation dataset.

NB: y-axis range changes in each plot.

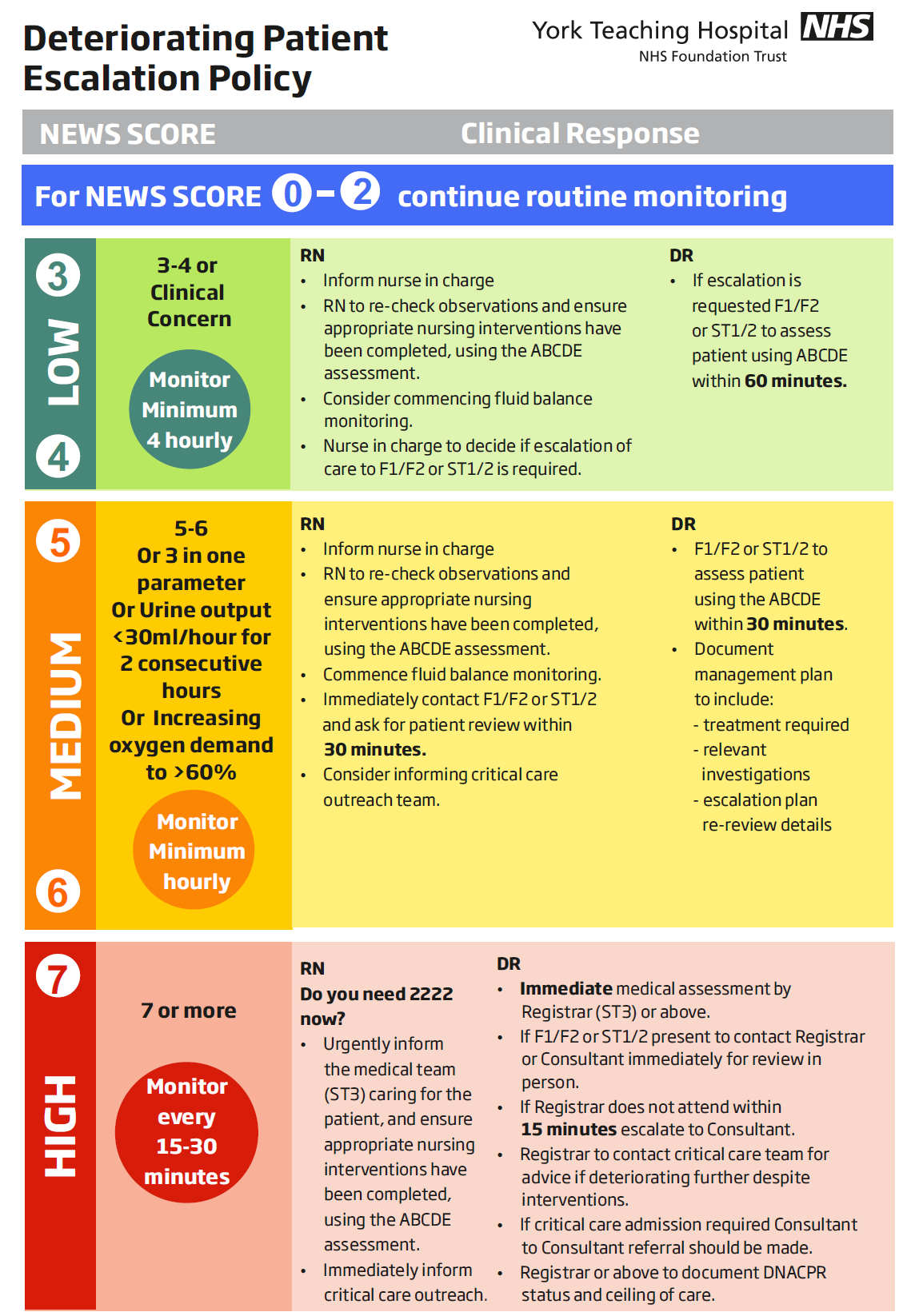

**Figure S5 Escalation policy of deteriorating patients in York Teaching Hospital NHS Foundation Trust.**

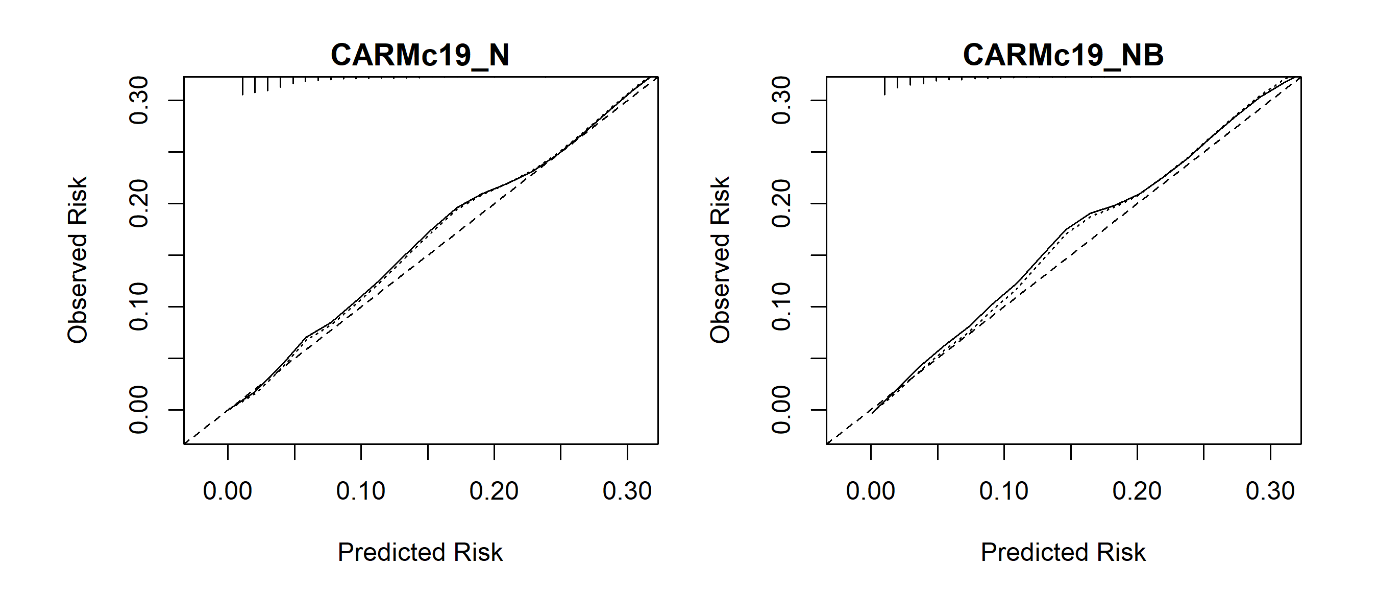

Figure S6 Calibration of CARMc19_N and CARMc19_NB models respectively for predicting the risk of mortality.

NB: We limit the risk of mortality to 0.30 for visualisation purpose because beyond this point, we have few patients.

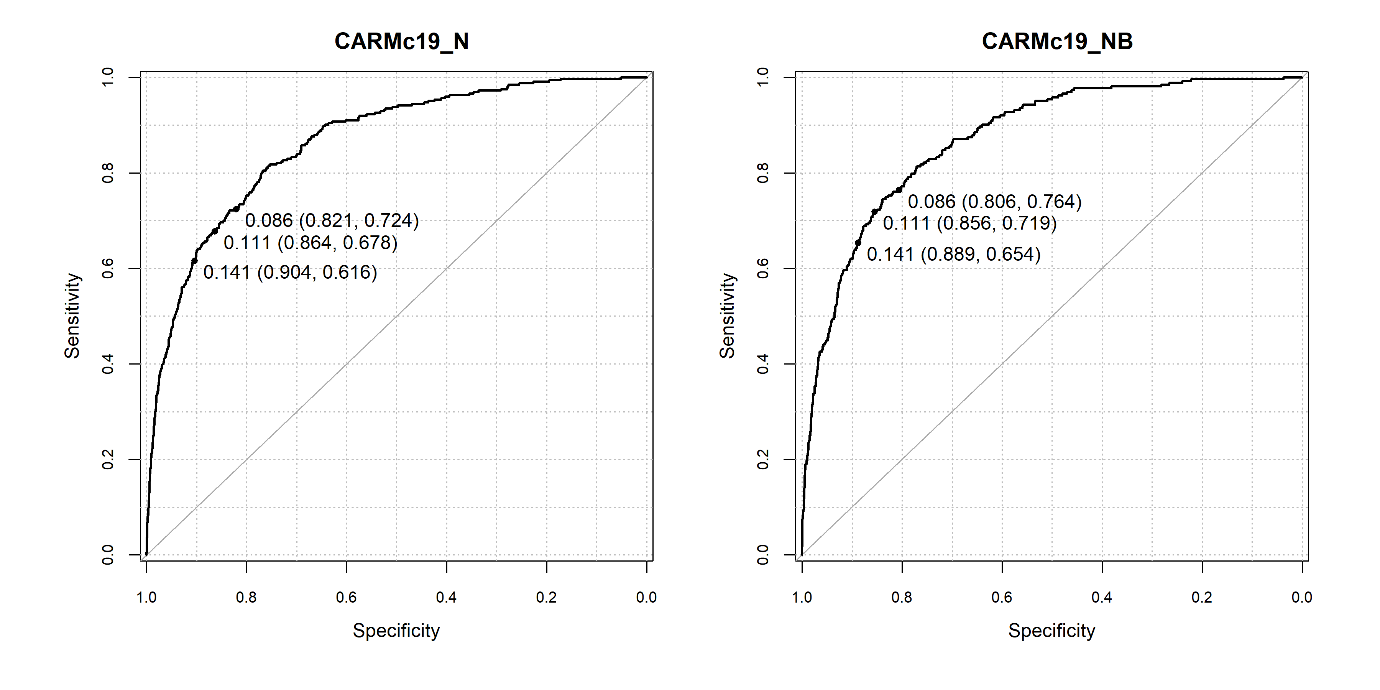

**Figure S7 Receiver Operating Characteristic curve for CARMc19_N and CARMc19_NB in predicting the risk of mortality in the development dataset.**

Note: predicted probability at NEWS2 thresholds 4+ [0.09], 5+[0.11], 6+[0.14] (sensitivity, specificity)

| **Model** | **Mean risk discharged alive** | **Mean risk discharged deceased** | **ARD** | **Scaled brier score** | **AUC (95% CIs)** | **Optimism-corrected c-statistic** |
| --- | --- | --- | --- | --- | --- | --- |
| CARMc19_N | 0.06 | 0.31 | 0.25 | 0.24 | 0.86  (0.84 to 0.87) | 0.85 |
| CARMc19_NB | 0.06 | 0.31 | 0.25 | 0.24 | 0.87  (0.85 to 0.89) | 0.86 |

**Table S3: Performance of CARMc19_N and CARMc19_NB models for predicting the risk of mortality in the development dataset.**

ARD: Absolute risk difference; AUC: Area under the curve; CIs: Confidence intervals

| **Model** | **At NEWS score [predicted risk of death]** | **Number of deaths identified by the model** | **Sensitivity%** | **Specificity%** | **PPV** | **NPV** | **LR+** | **LR-** |
| --- | --- | --- | --- | --- | --- | --- | --- | --- |
| CARMc19_N | 4+ [0.09] | 880 | 72.4  (67.2 to 77.2) | 82.1  (80.8 to 83.3) | 26.6  (23.7 to 29.6) | 97.1  (96.4 to 97.6) | 4  (3.7 to 4.4) | 0.3  (0.3 to 0.4) |
| CARMc19_N | 5+ [0.11] | 710 | 67.8  (62.4 to 72.9) | 86.4  (85.2 to 87.5) | 30.8  (27.5 to 34.4) | 96.8  (96.1 to 97.3) | 5  (4.4 to 5.6) | 0.4  (0.3 to 0.4) |
| CARMc19_N | 6+ [0.14] | 544 | 61.6  (56.1 to 66.9) | 90.4  (89.4 to 91.4) | 36.6  (32.5 to 40.8) | 96.3  (95.6 to 96.9) | 6.4  (5.6 to 7.3) | 0.4  (0.4 to 0.5) |
| CARMc19_NB | 4+ [0.09] | 750 | 76.4  (70.8 to 81.4) | 80.6  (79.1 to 82) | 26.4  (23.3 to 29.7) | 97.4  (96.7 to 98) | 3.9  (3.6 to 4.4) | 0.3  (0.2 to 0.4) |
| CARMc19_NB | 5+ [0.11] | 602 | 71.9  (66 to 77.2) | 85.6  (84.2 to 86.8) | 31.2  (27.5 to 35) | 97.1  (96.4 to 97.7) | 5  (4.4 to 5.6) | 0.3  (0.3 to 0.4) |
| CARMc19_NB | 6+ [0.14] | 496 | 65.4  (59.3 to 71.1) | 88.9  (87.6 to 90) | 34.8  (30.6 to 39.2) | 96.6  (95.8 to 97.2) | 5.9  (5.1 to 6.7) | 0.4  (0.3 to 0.5) |

**Table S4 Sensitivity analysis of CARMc19_N and CARMc19_NB models for predicting the risk of mortality at NEWS2 thresholds (4,5,6) for development dataset.**

PPV = Positive Predictive Value; NPV = Negative Predictive Value; LR+ = Positive Likelihood Ratio; LR- = Negative Likelihood Ratio
